# Homophily in risk and behavior complicate understanding the COVID-19 epidemic curve

**DOI:** 10.1101/2021.03.16.21253708

**Authors:** Nina H. Fefferman, Matthew J. Silk, Dana K. Pasquale, James Moody

## Abstract

New COVID-19 diagnoses have dropped faster than expected in the United States. Interpretations of the decrease have focused on changing factors (e.g. mask-wearing, vaccines, etc.), but predictive models largely ignore heterogeneity in behaviorally-driven exposure risks among distinct groups. We present a simplified compartmental model with differential mixing in two behaviorally distinct groups. We show how homophily in behavior, risk, and exposure can lead to early peaks and rapid declines that critically do not signal the end of the outbreak. Instead, higher exposure risk groups may more rapidly exhaust available susceptibles while the lower risk group are still in a (slower) growth phase of their outbreak curve. This simplified model demonstrates that complex incidence curves, such as those currently seen in the US, can be generated without changes to fundamental drivers of disease dynamics. Correct interpretation of incidence curves will be critical for policy decisions to effectively manage the pandemic.

## Introduction

Early winter saw a rapid and rise in incidence of COVID-19 leading to unprecedented levels of hospitalizations and deaths (*1*). Although many epidemiological models predicted that the US winter outbreak would peak in January, the case counts have dropped much more precipitously than anticipated in late winter (Figure 1). The wide swings in infection rates have led to broad pronouncements about causes (*2*), ranging from changes in social distancing policies (*3, 4*), seasonality (*5*), and disease strains (*6, 7*).

**Fig 1.**
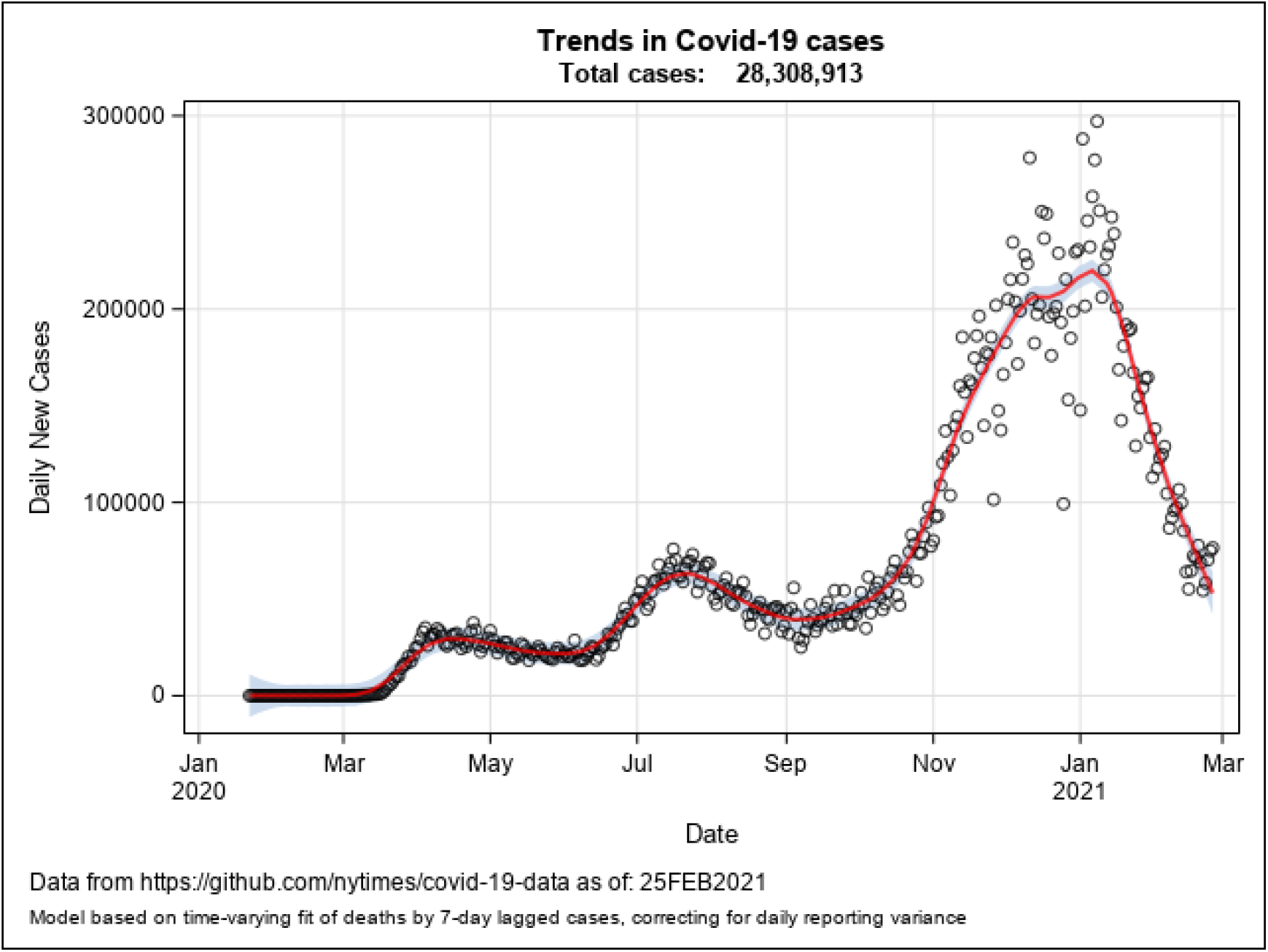
Trends in COVID-19 incidence in the US. Estimated daily new SARS-CoV-2 cases in the United States from January 2020 to March 2021.

Largely absent from such popular accounts is any recognition of the role of heterogenous population mixing and resulting differences in natural spread within groups, each with somewhat different epidemic thresholds, cumulative caseloads, and potential to approach (local) herd immunity. In a large, heterogenous population like the United States with wildly different approaches to masking, closures, and social distancing we would expect spread to be similarly uneven, though these sorts of features are often lost in national trends. Here, we present a simplified two-group model based on differential mixing that correlates with transmission risk (of course, real-world cases likely involve multiple populations, each with different behavior and risk profiles). This demonstration model is intended to highlight the simplest case for how homophily in behavior, risk, and contact makes compound epidemics with multiple “waves” likely by generating population(s) where the epidemic moves quickly loosely coupled with other population(s) where the epidemic creeps along much more slowly.

### Background

#### Explanations for wildly varying incidence rates

Naturally, decreases in case incidence under the complex and shifting realities of the pandemic are unlikely to be the result of any sole cause. The explanations already being offered are compelling and likely influence observed incidence. There are undeniable impacts on transmission from the timing of the winter holidays, including the rise and drop in travel (*8*), increased propensity to gather with family despite restrictions (*9*), and also increased willingness to be tested despite not yet experiencing being symptomatic to allow travel and family gathering (*10*). The last of these may have meaningfully altered how effective we were as a nation at interrupting asymptomatic spread for a small period of time. Given the recent estimates of how much transmission may be driven by asymptomatic infections (*11*), this by itself may have had a profound impact on curtailing community spread and contributed greatly to current decreases in ways also not being discussed.

There may also be a nontrivial impact of seasonality on the transmissibility of COVID-19 due to virological features, impacted either by temperature and humidity in both outdoor and HVAC-controlled environments or else to crowding patterns and similar behavioral changes (*12*). The increasing availability of vaccines, especially among those who may have acted as conduits for transmission (e.g. essential workforce) is undoubtedly beginning to limit spread somewhat. Even winter weather challenges in testing and reporting may contribute to decreased observability in cases, on top of actual reduction in disease incidence (*13*).

Beyond mixing changes due to the holidays or weather-related behavior, there are also likely ongoing gradual changes in local adoption rates of mask wearing or social distancing, especially as local case incidence created greater local awareness of potential disease severity than may have been believed before direct observable outcomes due to differences in national reporting and media consumption (*14-17*). These regional differences in behavior lead to differences in community vulnerability, causing a potential feedback loop between behavior and local outbreak severity (*18*). Certainly, there have been some areas of the country that have experienced such high prevalence that the number of individuals with natural immunity after recovery should now begin to slow transmission (*19*). This is especially true in regions where ongoing super-spreader facilities have raged uncontrolled, continually seeding additional infection into the broader community, such as jails, workplaces, congregate living environments, etc. (*20-24*).

While all of these features likely affect spread to one degree or another, people are remarkably consistent over time in who they interact with (with holidays being the obvious exception that proves the rule). As we show below, homophily in social contact can be sufficient in itself -- holding disease and context dynamics constant -- to generate widely varying disease incidence profiles.

#### Heterogenous social mixing

Social segregation due to homophily is probably the single most well-known feature governing American social contact. For both strong and weak ties, people tend to come into contact with those who are like themselves at much higher rates than those with whom they differ, across multiple dimensions (*25*). These patterns are reinforced by strong ethnic, educational, and geographic correlates in occupations (*26, 27*). While homophily causes social segregation, it can take remarkably little social contact to enable infectious disease to spread quickly across such boundaries, with super-spreader events or rare high-activity nodes sufficient to bridge populations (*28*).

Attribute homophily is generally well-correlated with behavior homophily, which in the context of contagious disease modeling means that we would expect people in close contact with each other to share general practices, cautions and behaviors relevant for disease spread. For example, mask wearing might become *de rigueur* in some neighborhoods while largely spurned in others for a variety of reasons (*29-31*). Similarly, people who work in high-contact settings at highest risk likely have friends, contacts and family members working in similar situations, themselves also at higher risk.

These simple baseline features of social contact networks imply heterogenous transmission dynamics -- close contact networks with risk-behavior homophily should lead to rapid spread within groups that take on “Dangerous” behavior profiles, while the disease should spread slowly in those with “Safer” profiles.

To illuminate how homophily in COVID-19 risk behavior can drive complex case dynamics, we used a simplified heterogenous social system with two relevant groups: a group of Dangerous actors who have high susceptibility and transmission rates and a group of Safer actors who have lower transmission rates (Figure 2). The system is then governed by three sets of parameters: (1) the relative size of the two populations, (2) differences in transmission rates within & between populations, (3) contact rates between populations.

**Fig 2.**
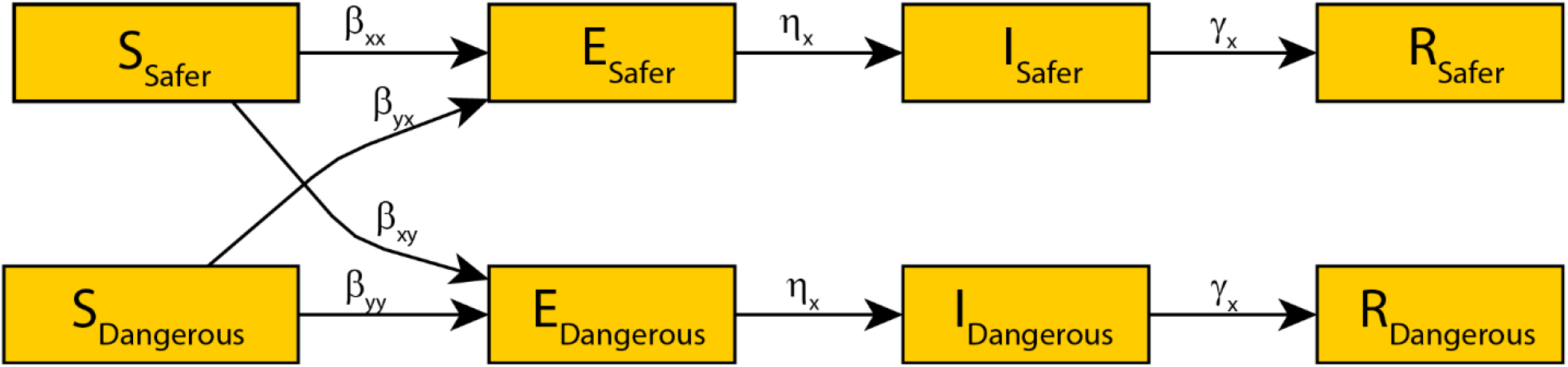
Simplified model schematic. A schematic of the simplified model of disease dynamics in two groups, one Safer with regard to behaviors affecting exposure risks and the other more Dangerous.

## Results

To illustrate how these dynamics work, we start with a simple baseline scenario, as specified in Table 1 (see Figure 3a). In our baseline scenario there is a faster infection rate for the Dangerous population than the Safer, but the two are highly correlated in time and build on each other smoothly, resulting in a single-phase total infection curve. We then consider differences in well-known health disparities by assuming that the populations (*x* and *y*) differ in underlying health risks and access to care, captured by a longer infectious period in the Dangerous population (0.8*\*γ*_*Safer*_) and higher death rates (1.2*\*ω*_*x*_). Here we again see a smooth overall transmission pattern with rapid Dangerous population spread (Fig. 3b).

**Table 1.** Values used in the Baseline Model presented in the main text. (For sensitivity of disease dynamics to these choices, see Supplementary Materials.)

| Parameter | Meaning | Value Used |
| --- | --- | --- |
| $\beta_{x,y}$ | Composite probability of contact and transmission between individuals in groups x and y | $\beta_{x,x} = 0.001$<br>$\beta_{x,y} = 20 \cdot \beta_{x,x}$<br>$\beta_{x,y} = \beta_{y,x} = \begin{cases} \beta_{x,x} & \text{under Baseline} \\ 0.01 \cdot \beta_{x,x} & \text{under Homophily} \end{cases}$<br>Where here, x is the Safer group and y is the Dangerous group |
| $\eta_x$ | The rate of developing symptoms after catching the disease | $\eta_x = \eta_y = 1/14$ |
| $\gamma_x$ | The rate of recovery from infection | $\gamma_x = \gamma_y = 1/10$ |
| $\omega_x$ | The rate of death from infection | $\omega_x = \omega_y = 1/1000$ |
| States | Initial Condition | Value Used |
| $S_x$ | $S_x = S_y$ | 499 |
| $I_x$ | $I_x = I_y$ | 1 |
| $E, R, \text{ and } D$ | $E_x = E_y, R_x = R_y, \text{ and } D_x = D_y$ | 0 |

**Fig 3.**
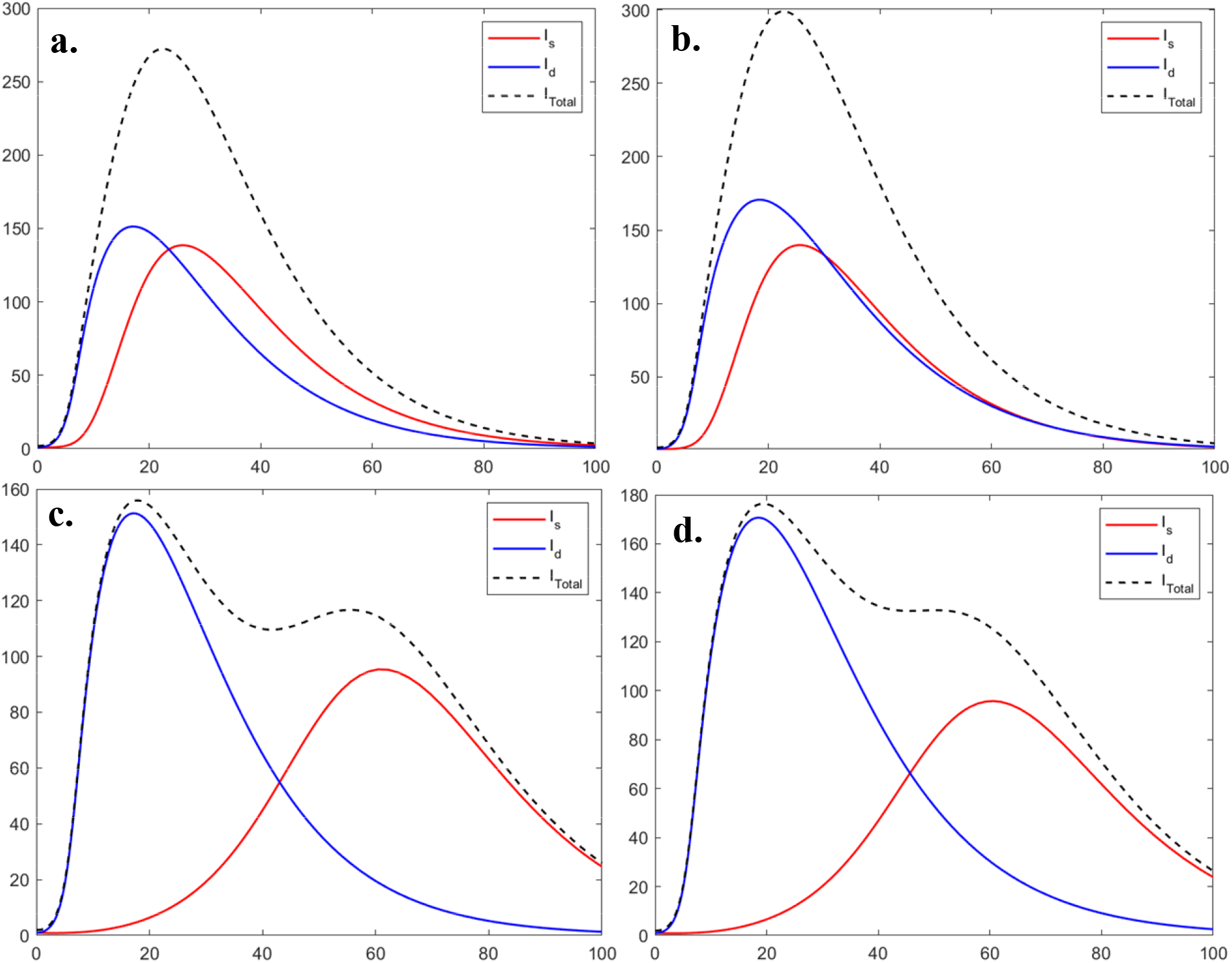
The impact of homophily and health disparities between the Safer and Dangerous Groups on observed case incidence. (**A**) The Baseline scenario with two groups that have different levels of behavioral exposure risk, but make contact across groups as often as within their group. (**B**) Building on the Baseline Scenario, the groups have uniform contact with each other, but now the Dangerous population also includes assumed increase in prevalence of underlying health conditions that increase the probability of severe infection outcomes. (**C**) Departing from the Baseline scenario, there is now homophily in group contact rates, rather than uniform contact across the groups. (**D**) Departing from the Baseline scenario, there are now both differential underlying health conditions and homophily.

If we now assume that mixing across populations is lower than mixing within populations, i.e. homophily such that *κ*_*x,Y*_ = 0.01 * *κ*_*x,x*_, we effectively allow spread to be much more highly compartmentalized (Fig. 3c). Adding homophily in this way leads to a classic “double hump” incidence curve with the Dangerous population leading infection early in the outbreak and a later, smaller peak in prevalence (slow burn outbreak) among the Safer population (see Figure 3c).

If we include both homophily and concomitant differential health and recovery risks for the Dangerous population, we see a less pronounced second “hump”, though there is still a critical plateau after the decrease from the initial peak as case incidence falls (Fig. 3d). In this particular case, the peak is lower and slightly earlier, but then transitions from an outbreak primarily among the Dangerous population into a “longer slow burn” outbreak among the Safer population.

The examples above are draws from a much wider space of potential models governed by the between-group differences in transmissibility and relative sizes of the two populations. We can examine the wider state space by sweeping a wide range of potential values (Fig. 4 and explored further in Supplementary Information).

**Fig 4.**
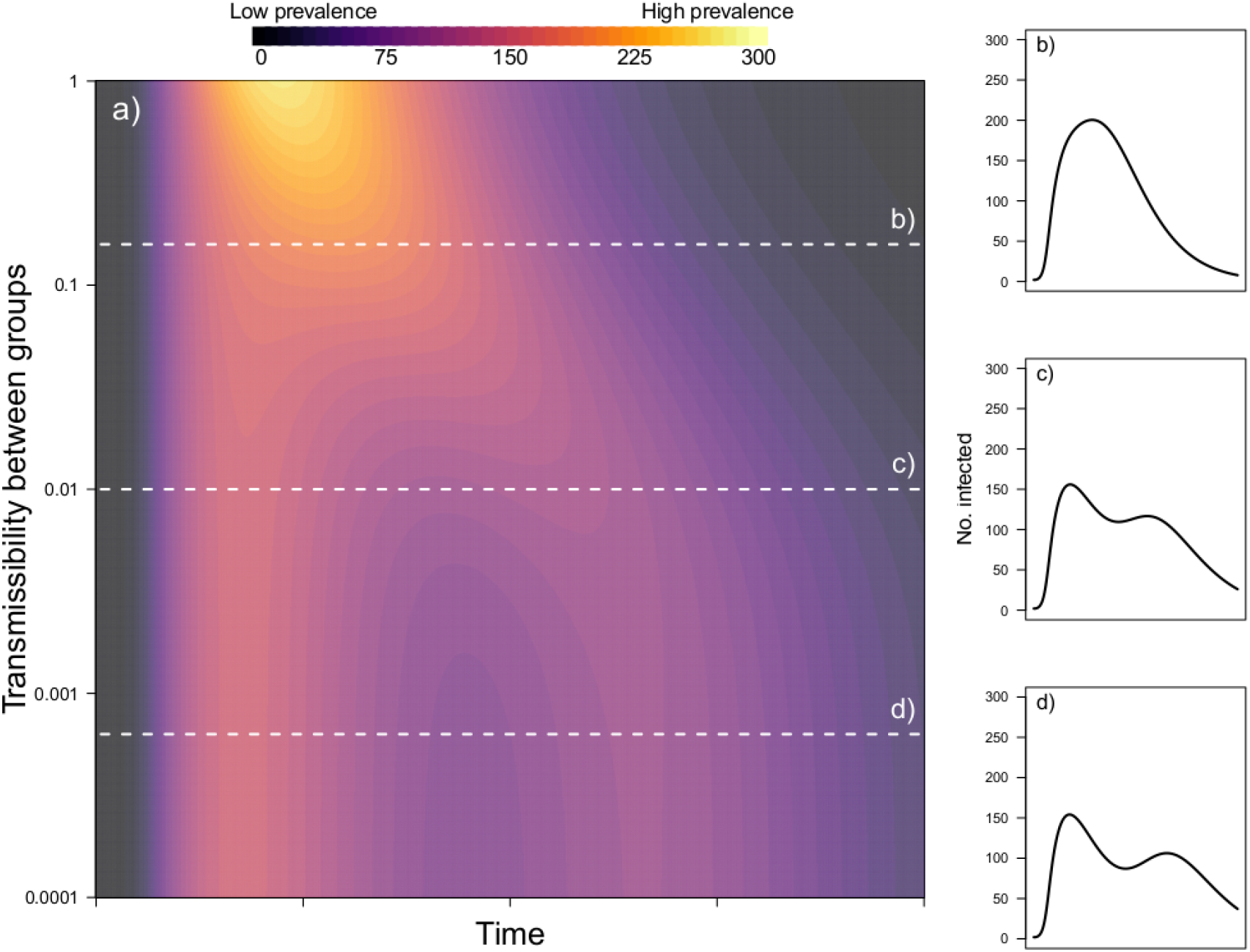
Varying between-group transmissibility. The effect of the difference of the between-group transmission rate (relative to the transmission rate in the Safer group) on epidemic curves when the transmission rate in the Dangerous group is 20 times that in the Safer group and each group makes up half the total population. Current prevalence in shown by color in panel (**A**) with panels (**B-D**) showing the epidemic curve for the parameter values indicated.

In Figure 4a we see that when transmission within groups is much higher than transmission between groups (low half of the figure), we get a quick outbreak among the Dangerous population that starts to burn out before the Safer population peaks, leading to a double-hump epidemic (Fig. 4d). As transmissibility between groups becomes more similar to transmission within groups this double-hump becomes of a plateau (Fig. 4c) and then is lost entirely (Fig. 4b).

The shape of the compound epidemic curve will also depend on the relative sizes of the different populations. In our case, a double-hump epidemic occurred when the Safer population was similar in size or bigger than the Dangerous population (Fig. 5). When a substantial majority of individuals were in the Safer population it was possible for the second epidemic peak of a double-hump epidemic to be higher (Fig. 5c).

**Fig 5.**
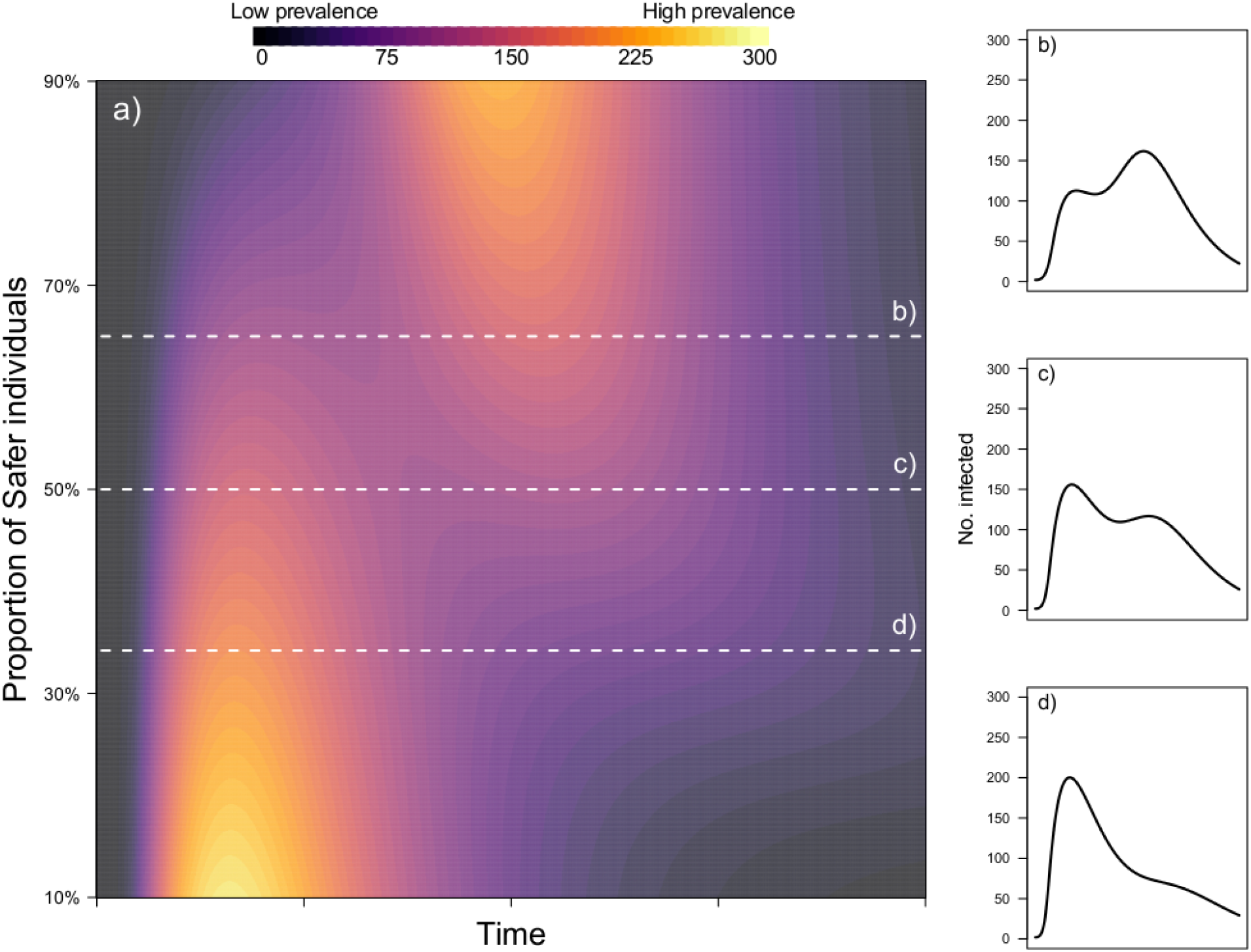
Varying relative group size. The effect of the relative sizes of Dangerous and Safer groups on epidemic curves. Current prevalence in shown by color in panel (**A**) with panels (**B-D**) showing the epidemic curve for the parameter values indicated.

One further complication in the observed incidence from aggregated populations with different health protective behaviors is that they may also have different rates of observability due to differences in testing access and willingness to be tested. Essential workers, for example, facing financial strain if diagnosed may have an incentive to avoid testing (*32*) or simply have little access to testing (*33-36*). The models thus far have assumed a transparent and complete disease detection system, however, to demonstrate this further potential confounder, we demonstrate one case that incorporates lower case detection rates within the Dangerous population.

The lower detection of course does not change the underlying spread pattern in these models, so this creates a situation where rapid, early spread goes somewhat unnoticed, but the spillover infections to other populations is detected at higher rates. This situation can be particularly problematic if the rise in infectivity among Safer populations could be attributed to new variants or other underlying disease dynamics, when in fact all we have is differential observability (Fig. 6).

**Fig 6.**
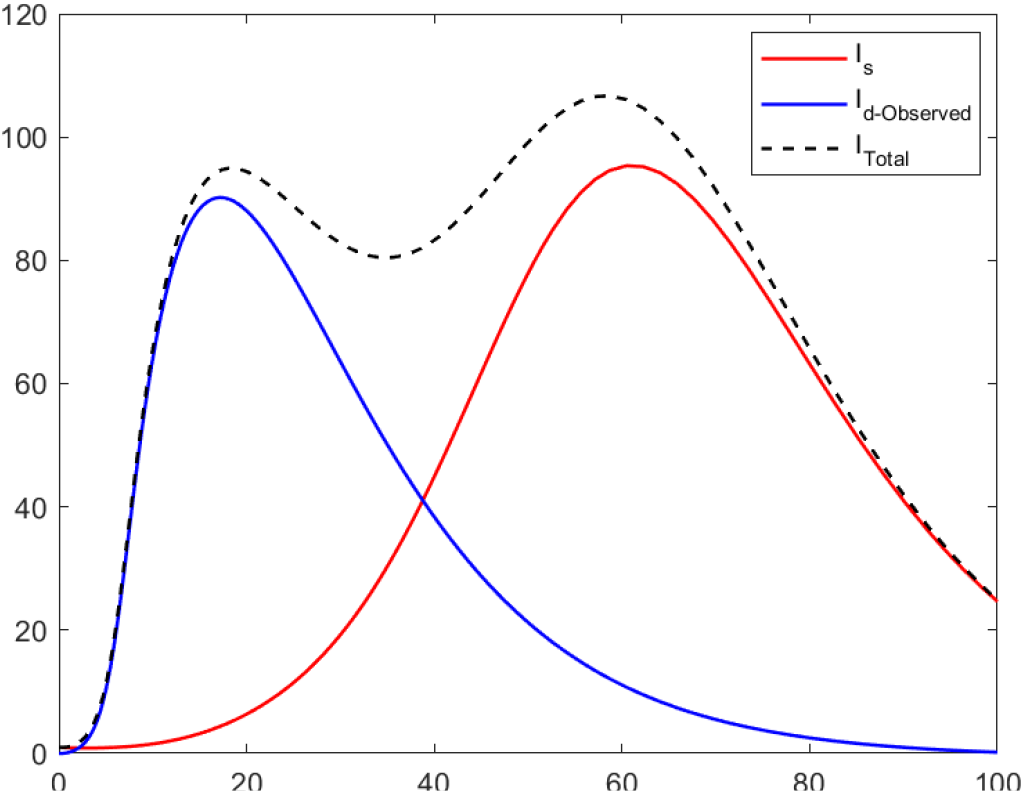
Differential observability. Observed incidence curves with high in-group homophily such that mixing across groups is lower than mixing within groups (equivalent to Figure 3c above) but also assuming that new cases of infection in individuals in the Dangerous group are only 60% as likely to be tested (and therefore reported) as individuals in the Safer group. While the infection dynamics are not altered by the observability of the cases, the surveillance curves are.

### Summary of results

Our simplified model demonstrates that myriad complex incidence curves can be generated without changes to fundamental disease dynamics or even overall changes in people’s everyday behavior. Rather, disease spread in America may simply reflect (at least in part) the underlying inequalities and social segregation of Americans’ daily activities. These interaction patterns govern who spreads disease to whom, who has access to healthcare and prevention tools, and who is likely to be tested. Hence inequalities can lead simultaneously to differences in the way the disease spreads and is observed.

## Discussion

Explanations for the current trends in spread have been characterized largely by exogenous features. Some of these include policies aimed at increasing social distancing or incentivizing masking and testing. Others focus on policy-independent extrinsic factors, such as seasonality in transmissibility due to climactic factors, or temporal patterns in travel and interaction rates within the US population due to holiday travel. While such features are certainly likely to be relevant, there is also a fundamental mixing assumption built into these explanations that should be examined. A simplified mixing model that distinguishes groups by risk can account for complex observed incidents curves without reference to radical policy, disease, or behavior change.

### Limitations of the study

Our goal is to demonstrate the potential impact of homophily on observed dynamics, rather than to make concrete quantitative predictions about actual reported incidence curves. Consequently, the model presented is only meant as a simplified example, and many potentially critical details are omitted, such as demographic or socioeconomic correlates with group behaviors. We also do not explicitly model the alternative explanations for the current trends. We do not mean to suggest that these factors are not playing a (large) role in the US COVID-19 pandemic, but aim to highlight a largely un-discussed, and potentially very important, additional influence of homophily among groups.

We have here demonstrated the impact of homophily in only two behaviorally distinct groups, though of course the reality is likely the composite contribution of many distinct groups that may vary in both behavior and/or physiological susceptibility (*37*). Sociocultural factors may also play a critical role in the rates at which different groups interact, since even beyond the percentage of households with economic or “essential workforce” constraints against protective behaviors, crowded neighborhoods and multigenerational homes also complicate the ability to minimize exposure risks (*38*). While heterogeneous mixing among such distinct etiological and behavioral cohorts is certainly not the only factor influencing currently observed trends, it would be a mistake to ignore the potential contribution of this effect going forwards. If these heterogeneities are sufficiently distinct, as they well may be (*39, 40*), the current rate of decline in new cases may easily slow, or even reverse, leading to additional future waves as an inevitable feature of social dynamics and disease behavior in the United States. Without including heterogeneous social mixing patterns into our considered factors, such dynamics may be attributed directly and completely to the gradual failures of previously successful mitigations, or to the emergence of new variants that are capable of overcoming available behavioral protections and/or vaccines. Health policy reactions to misattributed patterns could inadvertently abandon successful strategies, or even erode public trust and adherence without any shift in policy from leadership. It is therefore critically important to understand the contribution of such factors to potentially inevitable dynamics in patterns of incidence as the COVID-19 pandemic continues to unfold.

## Conclusions

Homophily between sub-populations that vary in their behavior and susceptibility to an infectious disease can drive compound epidemics with multiple waves even in the absence of behavioral change, seasonality, or antigenic escape. Consequently, correctly accounting for the impact of homophily will be vital in unpacking the success of non-pharmaceutical interventions or impact of new variants of concern, and so critical to forming effective health policy that retains broad political and public support.

## Materials and Methods

### Experimental design

#### SIR simulation modeling

There is a long tradition of using simplified mass-action compartmental models for disease spread (*41-43*), which are generally quite effective for diseases spread easily such as influenza or coronavirus related diseases. Such models assume subsets of a population can be divided into compartments with uniform transitions across disease states. Transitions between states are then governed by a set of transmission rates with the entire population system governed by a set of transmission rates. Using mass-action models to accurately fit for real-world data is complex, though versions are being used for short-term forecasting of the current pandemic (*44, 45*). However, simpler models remain effective as ways to build insights into the interdependencies of hypothesized system behavior, through building on a long social simulation tradition (*46, 47*).

#### A Behavior-correlated Two Population Mixing Model

The system is designed as a two-population Susceptible-Exposed-Infected-Recovered-Dead (SEIRD) compartmental model with rates across states governed by a set of ordinary differential equations.

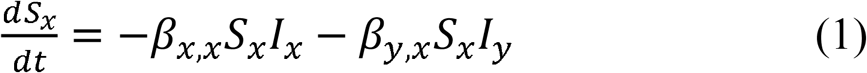

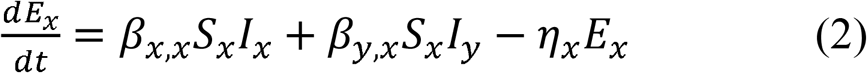

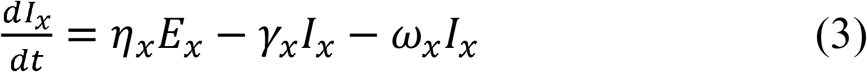

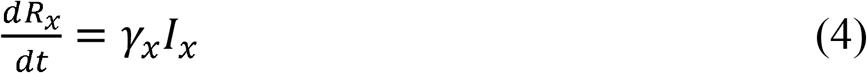

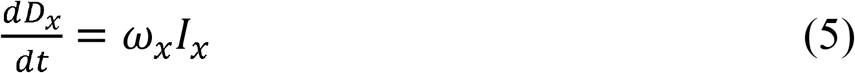

Where *β*_*x,Y*_ = *κ*_*x,Y*_ ·*min*(*λ*_*x,x*_, *λ*_*Y,Y*_) defined such that *κ*_*x,Y*_ is the probability of contact between an individual in population x and population y (the two groups), and *λ*_*x,x*_ is the probability of transmission given that contact occurs between two individuals within the same group, *x*. In this way, we assumed that transmission risks were dependent on the behavior of the individual who was adhering to the most disease protective behavior in any interaction. (For simplicity, this model did not include either infectious pre-symptomatic or asymptomatic individuals, though COVID-19 is known to involve such dynamics.)

The implementation of these equations was initially parameterized as the Baseline Model (see Table 1) and then altered as per case described in each reported scenario in the Results and Supplementary Materials.

## Data Availability

There is no primary data associated with this study.

## Funding

National Science Foundation award DEB 2028710 (NHF)

National Science Foundation award 2029790 (JM, DKP)

James S. McDonnell Foundation award 220020397 (JM)

## Author contributions

Conceptualization: NHF, JM

Methodology: NHF, JM, MJS

Investigation: NHF, MJS

Visualization: MJS

Writing—original draft: JM

Writing—review & editing: JM, NHF, MJS, DKP

## Competing interests

Authors declare that they have no competing interests.

## Data and materials availability

All data needed to evaluate the conclusions in the paper are present in the paper and/or the Supplementary Materials.

## Notes

### Competing Interest Statement

The authors have declared no competing interest.

### Funding Statement

The following funding supported this work, but did not influence study design or interpretation in any way:
National Science Foundation award DEB 2028710 (NHF)
National Science Foundation award 2029790 (JM, DKP)
James S. McDonnell Foundation award 220020397 (JM)

