## Supplemental Materials for "Homophily in risk and behavior complicate understanding the COVID-19 epidemic curve"

Supplementary Text

Explanation Supplementary Sensitivity Analysis

We tested the sensitivity of our two group mixing model by varying the values selected for three key social parameters: a) the proportion of Dangerous and Safer individuals in the overall population; b) the relative transmission rate of Dangerous versus Safer groups; and c) the between-group transmission rate (Dangerous-Safer/Safer-Dangerous) relative to the transmission rate among the Safer group.

a)	Group sizes: we varied the proportion of Safer individuals from 10% to 90% in increments of 0.1%

b)	Difference between groups: we varied transmission among the Dangerous group to be from 2 to 50 times greater than among the Safer group (800 increments).

c)	Between-group transmission: we varied the transmission rate between-groups to be between 0.0001 and 1 times that of the transmission rate in the Safer group (1000 increments on a log10 scale).

Supplementary figures here represent a sweep across this parameter space. Modelling and plotting was conducted in R 3.6.1.

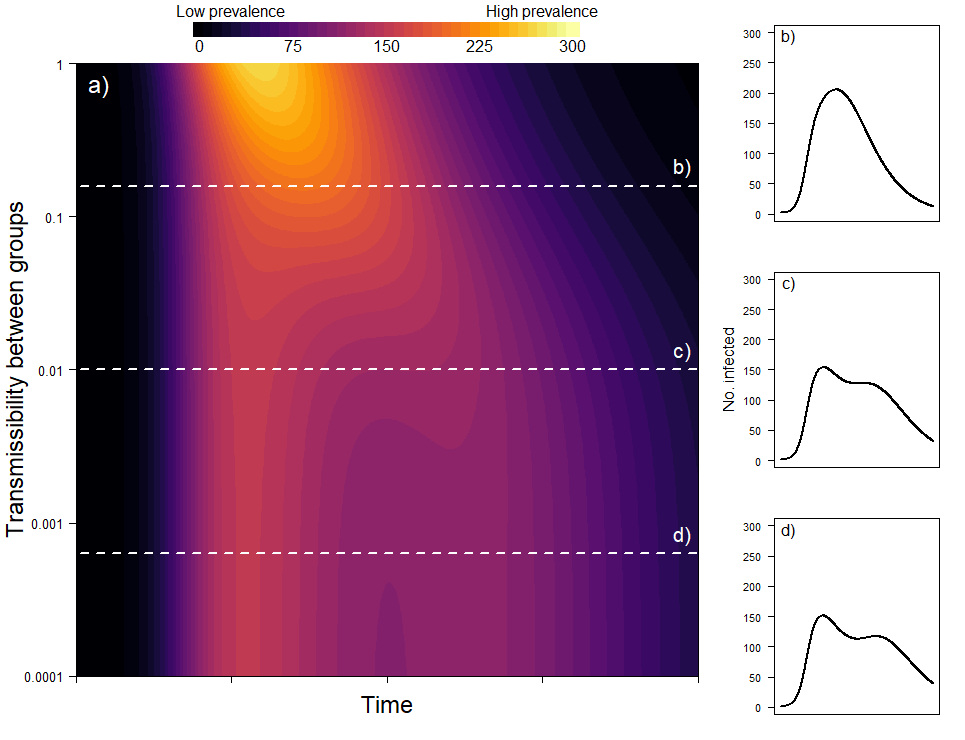

Fig. S1.

The effect of the difference of the between-group transmission rate (relative to the transmission rate in the Safer group) on epidemic curves when the transmission rate in the Dangerous group is five times that in the Safer group and each group makes up half the total population. Current prevalence in shown by color in panel a) with panels b-d) showing the epidemic curve for the parameter values indicated.

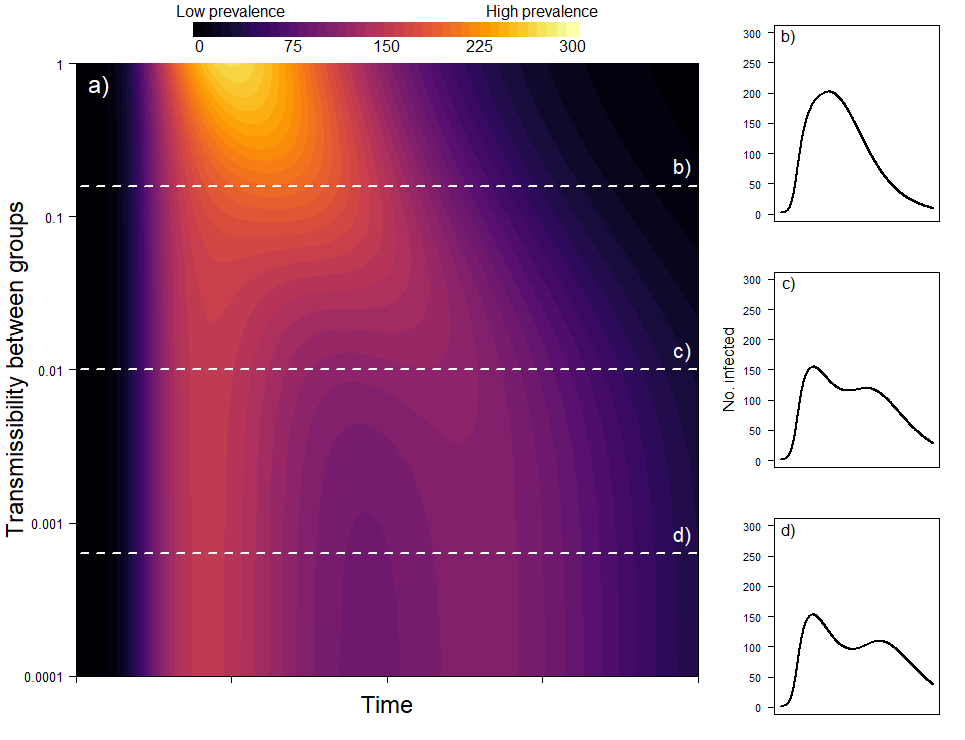

Fig. S2.

The effect of the difference of the between-group transmission rate (relative to the transmission rate in the Safer group) on epidemic curves when the transmission rate in the Dangerous group is ten times that in the Safer group and each group makes up half the total population. Current prevalence in shown by color in panel a) with panels b-d) showing the epidemic curve for the parameter values indicated.

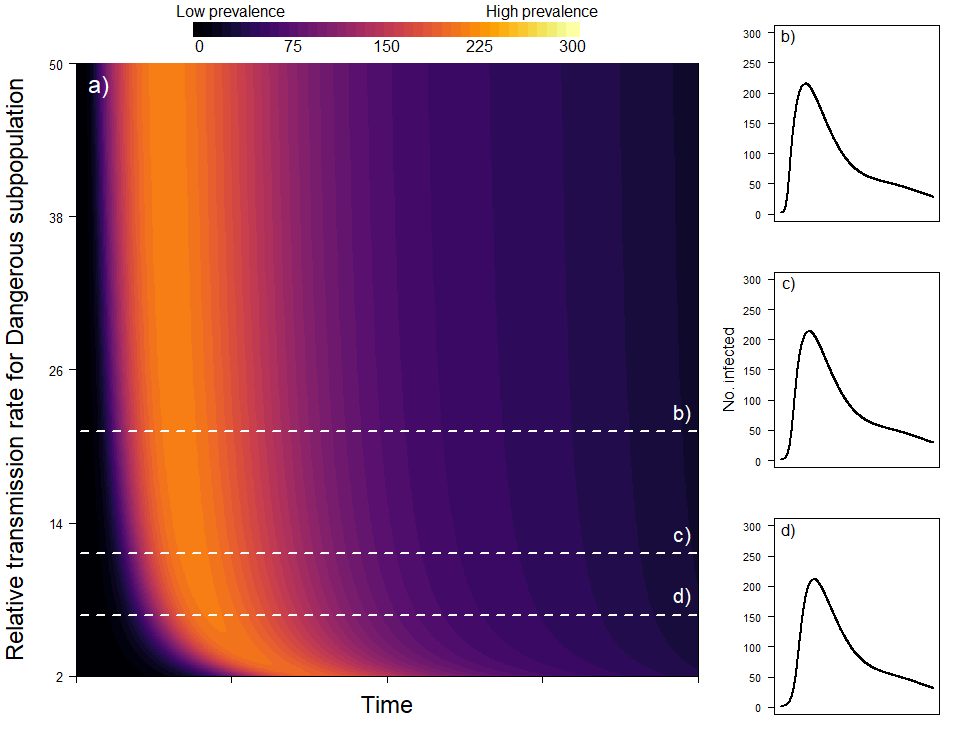

Fig. S3.

The effect of transmission rate in the Dangerous group (relative to the Safer group) on epidemic curves when the Safer group makes up 30% of the total population size. Between-group transmissibility is set to 0.01. Current prevalence in shown by color in panel a) with panels b-d) showing the epidemic curve for the parameter values indicated.

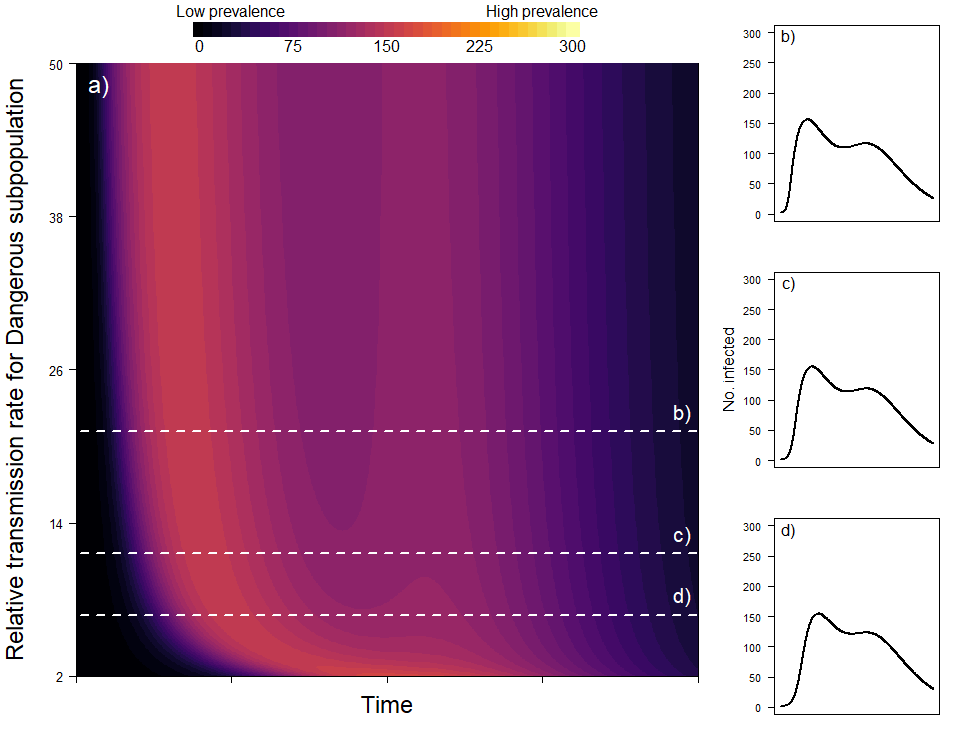

Fig. S4.

The effect of transmission rate in the Dangerous group (relative to the Safer group) on epidemic curves when the Safer group makes up 50% of the total population size. Between-group transmissibility is set to 0.01. Current prevalence in shown by color in panel a) with panels b-d) showing the epidemic curve for the parameter values indicated.

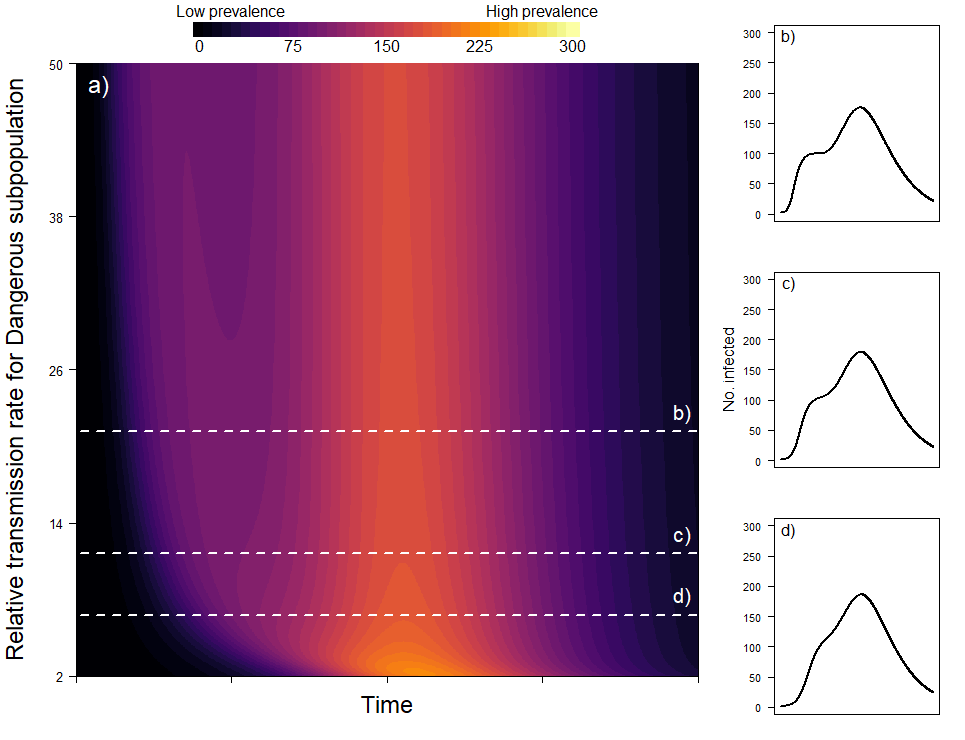

Fig. S5.

The effect of transmission rate in the Dangerous group (relative to the Safer group) on epidemic curves when the Safer group makes up 70% of the total population size. Between-group transmissibility is set to 0.01. Current prevalence in shown by color in panel a) with panels b-d)
